# Short report: A trend analysis of attitudes towards early diagnosis of dementia in Germany

**DOI:** 10.1101/2022.07.30.22278230

**Authors:** Andrea E. Zülke, Melanie Luppa, Tobias Luck, Steffi G. Riedel-Heller

## Abstract

**Background:** Early detection of dementia provides numerous benefits for those living with dementia and their relatives and healthcare systems at large. Methods available for early diagnosis have improved significantly over the past years. Therefore, we examined whether openness towards early diagnosis for dementia and preferred sources of support have changed in Germany over the last decade.

**Method:** We compared findings from two representative telephone surveys conducted among older adults in Germany (≥ 60 years of age) in 2011 and 2022, assessing attitudes towards and willingness to pursue an early diagnosis of dementia in a sample of n = 879 individuals (mean age: 72.9, % female: 58.8). Group comparisons using Chi^2^- and t-tests and multivariable regression analyses were conducted, assessing factors linked to willingness to be examined for an early diagnosis of dementia.

**Results:** Openness towards early diagnosis of dementia was high both in 2011 and 2022, but slightly declined over time (b=.62; 95% CI: .45; .86). Belief in preventability of dementia was linked to greater openness towards an early diagnosis of dementia (b=1.52; 95% CI: 1.12; 2.07).

**Conclusion:** Willingness to pursue an early diagnosis of dementia is high in the older German public, but slightly lower than reported previously. Improving knowledge on modifiable risk factors and better understanding of individual motives underlying endorsement or refusal of an early diagnosis may further increase acceptance in the general public.

## Background

Current estimates of people living with dementia are projected to increase from currently 55 million people worldwide up to 150 million in 2050 [1,2]. These estimates, however, only include those with a diagnosis of dementia. Diagnosis rates for dementia remain low, and globally, an estimated 75% of cases are undiagnosed, with numbers even higher in low- and middle-income countries [2].

With numbers of people living with dementia rising rapidly [3], importance of timely diagnosis of dementia has been emphasized by Alzheimer’s Disease International and the World Health Organization, among others [4–6]. This includes the use of biomarkers such as assessing hippocampal atrophy using magnetic resonance imaging or decreased levels of Aβ42 and elevated levels of tau and phosphor-tau in cerebrospinal fluid in preclinical stages of dementia [7,8].

Early diagnosis for dementia can entail several advantages. First, knowledge of the underlying condition can relieve uncertainty and irritation on the side of relatives and caregivers when early signs, e.g. forgetfulness or loss of orientation, become apparent, thereby facilitating adjustment and reducing emotional stress on the side of caregivers [9]. Knowing the reason for changes in cognition and behavior can further relieve stress on the side of those affected, as they no longer have to make efforts to hide difficulties related to impaired functioning in daily life [10]. Receiving an early diagnosis of dementia enables the person with dementia and his or her next ones to access care and make precautions for the future, e.g. arrangements of care and living situation, legal guardianship etc. [4,11]. Patient organizations like the German Alzheimer’s Association (Deutsche Alzheimer Gesellschaft) openly favor early visits to memory clinics or general practitioners (GPs) when noticing first symptoms of cognitive decline to pursue an early diagnosis of dementia [10].

On the other hand, ethical considerations including lack of available cure for dementia and the risk of psychological burden on the side of persons living with dementia, e.g. depressive symptoms, fear or suicidal ideation, are often raised as arguments against an early diagnosis [12]. Further, fear of losing control and independence, stigmatization, loss of social relationships and quality of life can raise fear on the side of older adults considering an early diagnosis of dementia [4,10].

For successful implementation in primary care, knowledge on acceptance and public attitudes towards early diagnosis of dementia is crucial. Acceptance of regular screening for Alzheimer’s disease in primary care has been reported high (71.2%) in a study conducted among older adults in Germany (60-80 years, mean age: 68.3 years; [13]. Similarly, a population-based survey conducted a decade ago among adults aged 18 years and older in Germany revealed high levels of support, with 87.9% in favor of offering a diagnosis and 68.6% willing to pursue a diagnosis themselves [14]. However, these studies were based on convenience sampling or conducted several years ago. Against this background, we investigated time trends in attitudes towards early diagnosis of dementia and preferred sources of professional help for dementia in the older general population in Germany. We therefore assessed openness towards early diagnosis of dementia and willingness to pursue a respective diagnosis in a population-based sample, applying the same methodology and assessments as used by Luck et al. [14] to allow for direct comparisons over time.

## Methods

### Recruitment and study population

We used data from two independent population-based surveys, conducted in 2011 and 2022, respectively. For both surveys, computer-assisted telephone interviews were conducted by USUMA GmbH, a German market and social research institute with expertise in health-related research. Participants were recruited applying a multi-stage random digit dialing procedure, using the Association of German Market and Social Research Agency’s (ADM) sample base, allowing for the inclusion of registered and non-registered telephone numbers from the German resident population. Telephone numbers were drawn proportionally to the respective German population structure in 2011 and 2022, stratified regionally according to district sizes. The Kish-Selection-Grid was applied within households to randomly select the target person to be interviewed within households (aged 18 or older in 2011 or 60 and older in 2022). A researcher from Leipzig University trained interviewers of USUMA GmbH for both surveys and interviewers were randomly monitored for quality control.

In the 2011-survey, 25,027 telephone numbers were contacted, identifying 5,897 eligible individuals. Among these, n=551 (9.3%) individuals could not be reached, refused to be interviewed (n=2,241, 38.0%) or discontinued the interview (n=102, 1.7%), leading to a sample of 3,003 complete telephone interviews. Questions on attitudes towards early diagnosis for dementia and preferred sources of support were asked in every third interview, therefor, the final analysis sample contained 1,002 individuals. For the 2022-survey, 1,067 individuals were initially selected. Excluding individuals who could not be reached (n=244, 22.9%) or refused participation (n=320, 30.0%), the final sample included n = 503 respondents. The two surveys included adults of different age groups (2011: adults aged 18 years and older; 2022: adults aged 60 years and older). To increase comparability, only adults aged ≥ 60 years in the 2011-survey were included in the present study. The final study sample included n = 879 individuals, n = 376 interviewed in 2011 and n = 503 interviewed in 2022.

### Instruments

For the present study, we used information assessed in both surveys using standardized questionnaires, i.e. participants’ sex, age, country of birth, education and occupational status (employed vs. retired, unemployed or homemaker). Education was assessed using the Comparative Analysis of Social Mobility in Industrial Nations (CASMIN)-classification, comprising information on professional and vocational education [15]. Participants were then asked about their support of offering an early diagnosis of dementia (“Do you think that early detection of dementia should be offered?”, response options: yes, no). Further, we assessed willingness to be examined for an early diagnosis (“Would you be willing to be examined for an early diagnosis of dementia?”, response options: in all cases, more likely, undecided, less likely, no). Participants’ belief in the preventability of dementia was assessed asking “Do you think that dementia could be prevented?” (response options: yes, no). Lastly, participants were asked about preferred sources of support for dementia diagnosis and care (“What would be your first source of professional help?”, response options: general practitioner, neurologist, psychiatrist, specialized services like memory clinics, other, I don’t know).

### Statistical analyses

Group comparisons were run using Chi^2^- and t-tests as appropriate. We assessed factors associated with willingness to pursue an early diagnosis of dementia using multivariable logistic regression analyses. Response options were dichotomized, with options “more likely” and “in all cases” indicating the will to be diagnosed for dementia, and options “undecided”, “less likely” and “no” indicating refusal. Willingness to pursue an early diagnosis for dementia was regressed on time, gender, age, education, employment status and belief that dementia could be prevented. All analyses were conducted using Stata 16.0 SE (StataCorp, College Station, TX, USA) and an alpha-level of 0.05 (two-tailed) was chosen to indicate significance.

## Results

### Sample description

**Table 1** presents sociodemographic information on the two samples, assessed in 2011 and 2022.

**Table 1:**
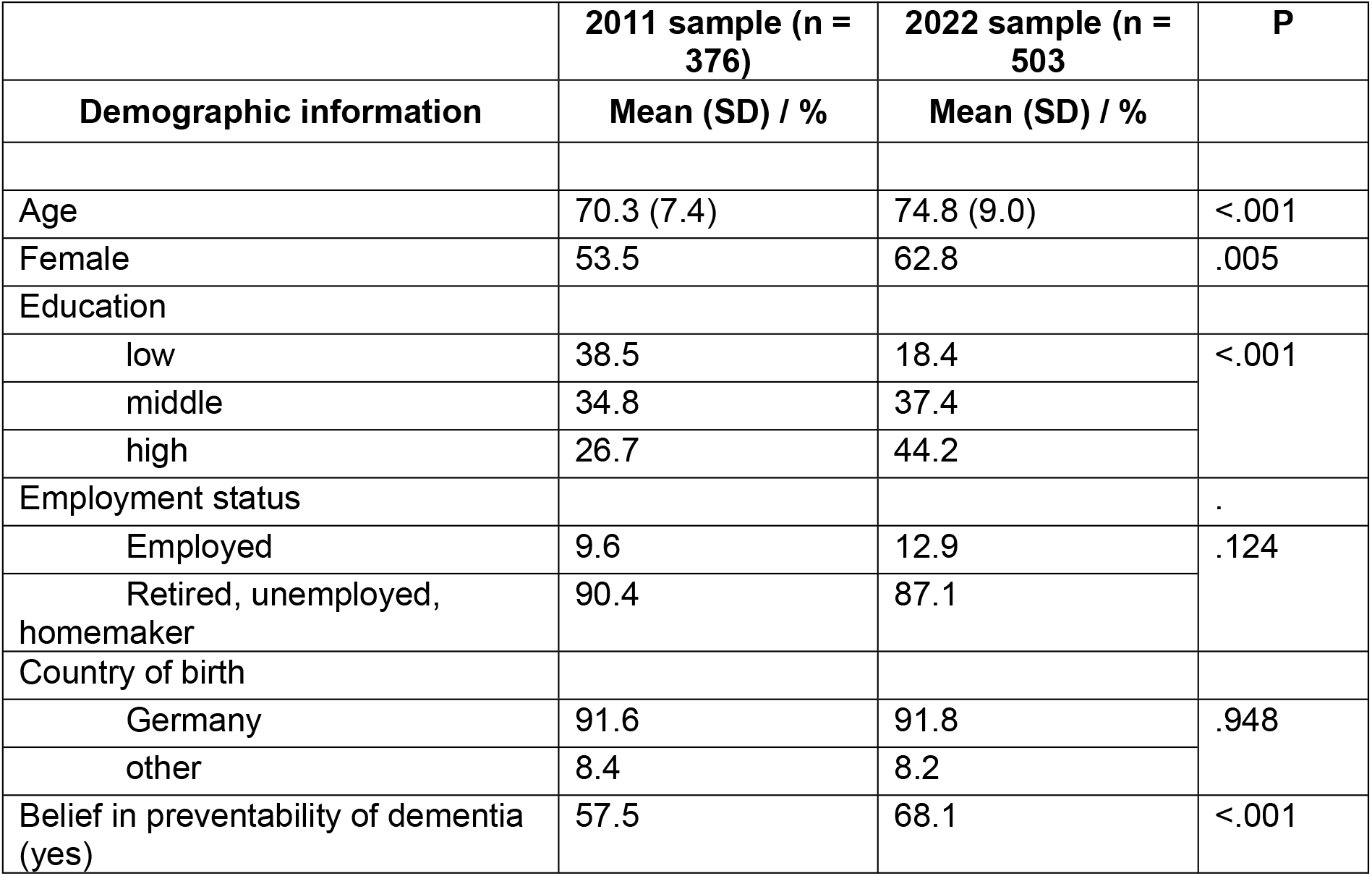
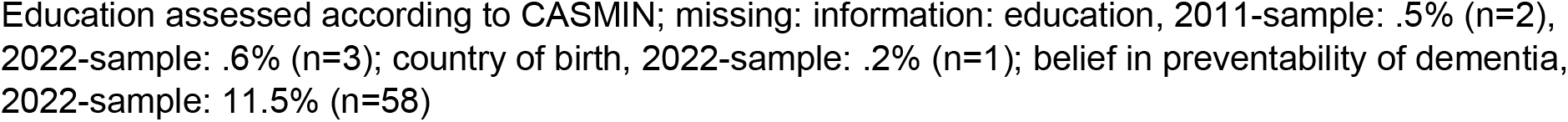
Sample description.

|  | 2011 sample (n = 376) | 2022 sample (n = 503) | P |
| --- | --- | --- | --- |
| Demographic information | Mean (SD) / % | Mean (SD) / % |  |
| Age | 70.3 (7.4) | 74.8 (9.0) | <.001 |
| Female | 53.5 | 62.8 | .005 |
| Education |  |  |  |
| low | 38.5 | 18.4 | <.001 |
| middle | 34.8 | 37.4 |  |
| high | 26.7 | 44.2 |  |
| Employment status |  |  | . |
| Employed | 9.6 | 12.9 | .124 |
| Retired, unemployed, homemaker | 90.4 | 87.1 |  |
| Country of birth |  |  |  |
| Germany | 91.6 | 91.8 | .948 |
| other | 8.4 | 8.2 |  |
| Belief in preventability of dementia (yes) | 57.5 | 68.1 | <.001 |

Education assessed according to CASMIN; missing: information: education, 2011-sample: .5% (n=2), 2022-sample: .6% (n=3); country of birth, 2022-sample: .2% (n=1); belief in preventability of dementia, 2022-sample: 11.5% (n=58)

Participants in the 2022-survey were more often female (p=.005) and had higher levels of education (p<.001) than participants in the 2011-survey. On average, participants in the 2022-survey were older than those participating in 2011 (p<.001). Belief that dementia can be prevented was higher in 2022 than in 2011 (p=.002). The two samples did not differ significantly with regard to employment status (p=.124) or country of birth (p=.948).

In both samples, participants named their attending GP as their preferred source of support regarding the condition (63.0% in 2011, 69.4% in 2022). Only 5.0% of participants in the 2022-survey preferred support provided by memory clinics, while this option was endorsed by 13.8% in 2011. Endorsement of other possible sources of support, i.e. neurologists (2011: 20.2%; 2022: 18.2%) or psychiatrists (2011: 1.4%, 2022: 2.4%) was comparable between the two surveys.

### Attitudes towards an early diagnosis of dementia

Support for offering an early diagnosis of dementia was high in both samples, but lower in 2022 than in 2011 (79.2% vs. 90.7%, **Table 2**). Asked whether they would be willing to pursue an early diagnosis of dementia themselves, participants in the 2022-survey less often endorsed the options “more likely” or “in all cases” than those in the 2011-survey.

**Table 2:** Attitudes towards an early diagnosis for dementia in 2011 and 2022.

|  | **2011 study** | **2022 study** |
| --- | --- | --- |
| **Support of early diagnosis for dementia** | n=364 | n=475 |
| Yes | 90.7 | 79.2 |
| No | 9.3 | 20.8 |
| **Willingness to be examined for an early diagnosis of dementia** | n=372 | n=499 |
| In all cases | 43.0 | 37.7 |
| More likely | 27.7 | 22.4 |
| Undecided | 9.1 | 13.2 |
| Less likely | 8.6 | 13.4 |
| No | 11.6 | 13.2 |

Factors linked to willingness to pursue an early diagnosis for dementia, assessed using multivariable logistic regression analyses, are provided in **Table 3**. In 2022, respondents less often stated wanting to pursue an early diagnosis of dementia (b = .62, 95% CI: .45; .86). The wish to pursue an early diagnosis was not linked to age, gender, education or employment status. Believing that dementia can be prevented was associated with the wish to pursue an early diagnosis of dementia (b= 1.52, 95% CI: 1.12; 2.07).

**Table 3:** Support of early diagnosis for dementia, regressed on sociodemographic characteristics and time (n=809)

|  | **Coeff.** | **SE** | **95% CI** | **P** |
| --- | --- | --- | --- | --- |
| Age | .99 | .01 | .97; 1.01 | .187 |
| Female gender | .77 | .12 | .56; 1.05 | .100 |
| Education (ref.: low) |  |  |  |  |
| middle | 1.22 | .24 | .84; 1.79 | .300 |
| high | 1.31 | .26 | .88; 1.94 | .183 |
| Employed (ref: retired, unemployed, homemaker) | .90 | .23 | .55; 1.49 | .691 |
| Belief in preventability of dementia (ref: no) | **1.52** | .24 | 1.12; 2.07 | .007 |
| Time (ref: 2011) | **.62** | .10 | .45; .86 | .005 |

CI: confidence interval; coeff.: coefficient; ref.: reference; SE: standard error; education assessed according to CASMIN; results present unstandardized beta-coefficients; significant associations in bold type

## Discussion

Our primary aim was to assess time trends in attitudes towards early diagnosis of dementia in the older general population in Germany, drawing upon data from two large population-based surveys conducted in 2011 and 2022 using the same methodology and questionnaires.

A vast majority of participants was in favor of offering an early diagnosis for dementia and willing to be examined respectively, however, the respective proportion has declined from 2011 to 2022. Our results might point towards greater fear of or skepticism towards the benefits of early diagnosis than reported a decade ago. The wish to pursue an early diagnosis of dementia or refusal thereof was not explained by differences in age, sex, education or employment status, as indicated by multivariable regression analyses. Our findings partly differ from a previous study from Germany, reporting that higher age was linked to greater skepticism towards an early diagnosis for dementia [16]. However, believing that dementia can be prevented was linked to greater openness towards an early diagnosis of dementia. Similar findings were reported by Hajek and König, who found that belief in preventability was linked to less fear of dementia in a population-based German sample of older adults [17].

Our findings suggest that strengthening knowledge on modifiable risk factors for dementia and opportunities to prevent or delay disease onset might increase openness towards being diagnosed for dementia. Further analyses revealed that support for an early diagnosis of dementia was linked to better knowledge of risk and protective factors for disease in the 2022-sample, and better knowledge of risk and protective factors predicted greater openness towards interventions for brain health (Zuelke et al., results submitted). Providing older adults with information and guidelines on how to preserve cognitive function in older age by means of a healthy lifestyle (e.g. management of cardiovascular risk factors, physical, social and cognitive activity, healthy diet) might strengthen belief that cognitive decline and dementia can be subject to intervention and preventive approaches. Respective efforts might in turn foster acceptance of early diagnosis of the condition. On the other hand, recent studies have identified a shift in public and medical discourses about dementia, with strong focus on individual means of risk modification, running the risk of shifting responsibility for the condition one-sidedly on the individuals affected [18,19]. Over-emphasizing the potential of prevention might lead to increased fear or shame in older adults if they experience signs of cognitive decline, possibly making them more reluctant to be examined for an early diagnosis of dementia. Since, however, we did not directly assess fear of dementia or perceived individual disease risk in our study, this line of thought should be interpreted with caution. Future studies assessing reasons for endorsement or refusal of early diagnosis of dementia in older adults are highly warranted. These could additionally assess further covariates possibly linked to attitudes towards an early diagnosis of dementia, e.g. perceived impact of a diagnosis on one’s future, beliefs about and attitudes towards people living with dementia.

Most older adults in our samples would prefer their attending GP as professional helper regarding dementia, with similar estimates across time. Only small numbers of respondents named other professions, e.g. neurologists, as preferred sources of professional help. This is in line with findings from several national and multinational studies, highlighting that GPs often constitute the first contact of patients regarding dementia diagnosis and care [20–22]. As most older people see their attending GP on a regular basis, often over extensive periods of time, GPs play an important role in detecting first signs of dementia and informing patients and relatives on further steps to be taken regarding treatment and care [23], as emphasized in the German National Dementia Strategy (“Nationale Demenzstrategie”; [24]). Our findings therefore illustrate the importance of enabling GPs to detect early signs of dementia in primary care and to inform people living with dementia and their families about further medical and legal steps to be taken. However, the small proportion of older adults naming memory clinics as preferred sources of support demands attention. Although the network of memory clinics as specialized institutions for the diagnosis of dementia and prodromal stages of disease has been expanding rapidly during the last decade, the proportion of older adults considering visits to a memory clinics declined by more than 50% in our study. Future studies are warranted to investigate awareness of memory clinics and their respective services and reasons for acceptance or refusal in the older general population in Germany. Respective investigations might inform actors in health policy and health care systems to make the best use of available resources in the long run.

### Strengths and limitations

Certain limitations need to be considered when interpreting our findings. We were not able to assess possible associations of marital status with openness towards early diagnosis of dementia, as this information was not collected during the 2011-survey. Caring roles and responsibilities for family members, especially spouses or partners, might likely influence attitudes towards provision and use of early diagnosis for dementia. Moreover, we cannot make statements about possible reasons underlying approval or refusal of early diagnosis of dementia as this information was not assessed in the two surveys. Questions assessing e.g. fear of dementia might provide further insights on why older adults endorse or refuse an early diagnosis of dementia.

This study uses data from two population-based surveys conducted using the same questionnaires and sampling technique, allowing for the investigation of time trends regarding attitudes towards early diagnosis of dementia in the German general population. Our findings might therefore provide insights that are more generalizable to different populations than e.g. studies using convenience sampling. Further, the use of two independent samples should make our results more robust against non-random sample attrition and selection effects often encountered in panel data studies [25]. Despite advances in available means for early detection of dementia and development of risk assessment tools, evidence on their acceptance in the general public is currently rare [26]. Knowledge on acceptance of early diagnosis of dementia in the general public is of critical importance, as approval of screening procedures might likely influence processes of decision making and improve adherence to agreements regarding treatment and care in older adults [27].

## Conclusion

By including data from a survey conducted a decade ago, we were able to identify trends in knowledge of preventability of dementia and attitudes towards early diagnosis for the condition in the older general population. Acceptance of and willingness to pursue an early diagnosis for dementia remain high in older German adults, although with slight decreases during the last decade. Improving knowledge on modifiable risk and protective factors for dementia, but also further investigations on what motivates older adults to endorse or refuse a respective diagnosis might help increase public acceptance of early diagnosis for dementia. Nevertheless, ethical considerations regarding dementia diagnosis and disclosure of results as well as different needs and preferences of older adults need to be taken into account when discussing means of early diagnosis of dementia.

## Data Availability

Data described in the manuscript will be made available at the Figshare repository upon acceptance.

n.a.

## Declarations

### Conflict of interest

The authors declare that they have no competing interests.

### Availability of data and materials

Data described in this article are available from the corresponding author upon request.

## Acknowledgement

We acknowledge support from Leipzig University for Open Access Publishing.

## Funding

This work was supported by a junior research grant awarded to AZ by the Medical Faculty of the University of Leipzig. The sponsor had no role in the design of the study or the collection, interpretation and presentation of data or decision to publish results.

## Authors’ contributions

AZ acquired funding for the study. AZ, ML and SR-H designed the study and supervised the data collection. AZ supervised the project and was supported by ML and SR-H. AZ analyzed the data and wrote the first draft of the article. ML, TL and SR-H revised the article for important intellectual content. All authors agree with the final version of the article.

## Ethics declaration and consent to participate

Our study was carried out in accordance with the principles of the Declaration of Helsinki in its revised version from 2000. The Ethics Committee of the Medical Faculty of the University of Leipzig approved the study (ref.: 587/21-ek). Interviewers informed participants verbally about the study at the beginning of the telephone interviews. Participants then provided oral consent, documented electronically by USUMA GmbH.

## Consent for publication

Not applicable.

## Notes

### Competing Interest Statement

The authors have declared no competing interest.

### Clinical Trial

n.a.

### Clinical Protocols

n.a.

